# Duration of Viral Clearance in IDF Soldiers with Mild COVID-19

**DOI:** 10.1101/2020.05.28.20116145

**Authors:** Tomer Talmy, Adili Tsur, Or Shabtay

## Abstract

Policies determining the duration of quarantine and return to work for confirmed COVID-19 patients still lack evidence. We report our findings regarding the duration of viral RNA positivity among a cohort of young patients with mild disease. Between March 20th, 2020, and May 10th, 2020, 219 soldiers were admitted to the Israel Defense Forces Medical Corps (IDF-MC) COVID-19 rehabilitation center following a positive RT-PCR test for SARS-CoV-2. 119 of these patients, 84 (70.6%) males, 35 (29.4%) females with a median age of 21 (IQR 19-25) were classified as having mild disease and had two consecutive negative RT-PCR tests by May 10th, 2020. The median time for SARS-CoV-2 positivity in nasopharyngeal or oropharyngeal swabs in the study population was 21 days (IQR 15-27) from symptom onset, with a range of 4 to 45 days. The results of this study suggest that in young and healthy adult patients with COVID-19, the median duration of viral positivity is around three weeks. This duration is higher than previously reported in other populations. Young and healthy adults comprise much of the population workforce, and the results of this study may assist in determining the isolation period for symptomatic adults and confirmed COVID-19 patients with mild symptoms. Further studies on this topic should look to expand and determine the intervals of serial testing for confirmed patients and determine the duration of SARS-CoV-2 positivity in other populations.

## Background

While considerable clinical and epidemiological data have been published during the 2019 novel coronavirus disease (COVID-19) pandemic,^1^ public health policies determining the duration quarantine and return to work for confirmed patients are still hampered by a lack of evidence. Real-time reverse transcriptase-polymerase chain reaction (RT-PCR) of upper respiratory specimens remains the primary detection method for COVID-19.^2–4^ Initial studies have reported a median duration of SARS-CoV-2 viral RNA positivity ranging 9.5–20 days from symptom onset^5–7^ and a median duration of two days in seven asymptomatic patients.^6^ An additional study found that 90% of patients with mild disease achieved viral clearance by ten days from symptom onset.^3^ We report our findings regarding viral RNA positivity duration among a cohort of young patients with mild disease.

## Materials and Methods

### Study Population

Between March 20^th^, 2020, and May 10^th^, 2020, 219 soldiers were admitted to the Israel Defense Forces Medical Corps (IDF-MC) COVID-19 rehabilitation center following a positive RT-PCR test for SARS-CoV-2, acquired from an oropharyngeal or nasopharyngeal swab. Recovery was defined as two sequential negative RT-PCR results, after which soldiers were discharged. We retrospectively reviewed patient clinical records and RT-PCR results to determine the duration of SARS-CoV-2 positivity, defined as the period between symptom onset and the first of two consecutive negative RT-PCR tests. For this study, we included recovered patients who presented mild symptoms and excluded patients who were asymptomatic, developed moderate disease or were lost to follow-up (Criteria available in the Supplementary Appendix).

### Data Collection

The medical staff at the rehabilitation center made telephone contact with patients upon receiving the initial positive RT-PCR results. Patient demographics, epidemiology, medical history, and symptoms were gathered upon initial contact and documented in the IDF Computerized Patient Records (CPR), and a separate database created for monitoring admitted patients. Upon admission to the rehabilitation center, patients filled an additional online questionnaire detailing demographic data along with current symptoms and vitals. During their stay at the rehabilitation center, monitoring physicians performed and documented daily telephone follow-up in an attempt to investigate symptom progression identify patients requiring further care. Patients who developed complications or experienced significant disease progression were referred to local tertiary hospitals for further care (Supplementary index).

### Specimen Collection and Viral RNA Detection of SARS-CoV-2

Nasopharyngeal and oropharyngeal swabs were obtained by specially trained IDF medics, paramedics or medical school students. Nasopharyngeal and oropharyngeal swabs were obtained within 48 hours of being asymptomatic. In the case of a negative result, an additional swab was obtained within 48 hours. If a positive result followed a negative one, additional swabs were obtained until two consecutive negative results were received.

### Definitions

Recovery was defined as two sequential negative RT-PCR results, after which admitted soldiers were discharged. The primary outcome of time to viral RNA clearance was defined as the time period between symptom onset and the first of two consecutive negative RT-PCR tests. This duration was then calculated with stratification for age, sex, and reported symptoms.

### Statistical Analysis

Continuous variables are described as medians with interquartile ranges (IQR) or means with standard deviations as deemed appropriate. Dichotomous variables such as the presence of symptoms are expressed as percentages. The date was aggregated in a dedicated spreadsheet and deidentified for the purpose of statistical analysis. The duration of SARS-CoV-2 positivity was assessed using Kaplan-Meier curves. The data were analyzed using IBM SPSS version 25.0 (Armonk, NY: IBM Corp).

### Ethical Considerations

The study was approved by the IDF-MC institutional ethics committee, which waived the requirement for informed consent.

## Results

Between March 20^th^, 2020 and May 10^th^, 2020, 119 symptomatic patients with two consecutive negative swabs were admitted to the IDF COVID-19 rehabilitation center, 84 (70.6%) males, 35 (29.4%) females with a median age of 21 (IQR 19–25). Symptoms reported upon admission included cough (75 [63.0%]), loss of smell (61 [51.3%]), headache (48 [40.3%]), loss of taste (47 [39.5%]) and fever (45 [37.8%]). 67 patients (56.36%) did not have any notable medical history (Supplementary Appendix). The clinical characteristics of the study population are detailed in Table 1.

**Table 1.** Patient characteristics, symptoms, duration of SARS-CoV2 positivity and patient comorbidities.

| Total ( N= 119) |  |  |
| --- | --- | --- |
| Age | Median (IQR), years |  |
| All patients (N=81) | 21 (19-25) |  |
| Sex | Number of patients (%) | Median duration of viral detection from symptom onset (IQR), days |
| Male (N, %) | 84 (70.6%) | 20 (15-27.75) |
| Female (N, %) | 35 (29.4%) | 21 (15-25) |
| Symptom upon admission | Number of patients (%) | Median duration of viral detection from symptom onset (IQR), days |
| All symptoms | 119 (100%) | 21 (15-27) |
| Cough | 75 (63.0%) | 21 (14-27) |
| Loss of smell | 61 (51.3%) | 21 (15-27.5) |
| Headache | 48 (40.3%) | 21 (16-25.75) |
| Loss of taste | 47 (39.5%) | 23 (15-28) |
| Fever | 45 (37.8%) | 19 (15-25.5) |
| Generalized Weakness | 41 (34.5%) | 20 (15-30) |
| Sore throat | 32 (26.9%) | 20.5(13-24.75) |
| Rhinitis | 30 (25.2%) | 20.5 (12.75-29.25) |
| Myalgia | 28 (23.5%) | 24 (17-33) |
| Dyspnea | 23 (19.3%) | 15 (12-24) |
| Diarrhea | 22 (18.5%) | 24 (17.75-29.25) |
| Loss of appetite | 16 (13.4%) | 23.5 (19.25-29.5) |
| Nausea | 11 (9.2%) | 26 (16-33) |
| Chills | 8 (6.7%) | 14 (12.25-23) |
| Abdominal Pain | 7 (5.9%) | 25 (21-32) |
| Chest Pain | 7 (5.9%) | 21 (15-24) |
| Rash | 5 (4.2%) | 27 (18.5-35.5) |

The median time for SARS-CoV-2 positivity in nasopharyngeal or oropharyngeal swabs in the study population was 21 days (IQR 15–27) from symptom onset, with a range of 4 to 45 days (Figure 1). The median time for viral RNA clearance stratified according to presenting symptom is detailed in Table 1.

**Figure 1.**
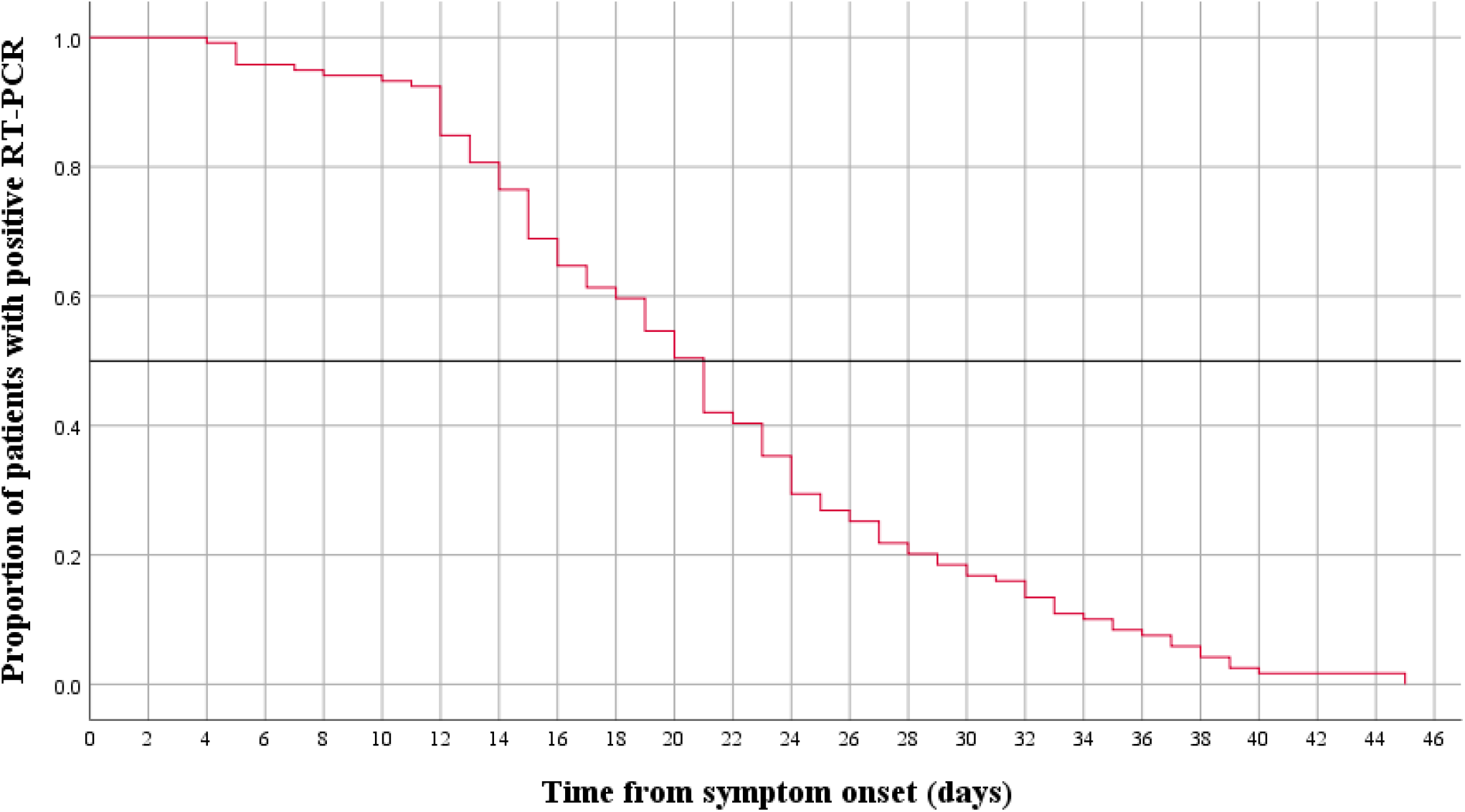
Kaplan-Meier curve of duration of SARS-CoV-2 positivity from symptom onset.

## Discussion

Determining the viral dynamics and natural history of SARS-CoV-2 in different populations is crucial towards instituting public health policies and granting clearance for infected patients. The results of this study suggest that in young and healthy adult patients with COVID-19, the median duration of viral positivity is around three weeks. This duration is higher than previous reports of a median between 9.5 and 20 days.^5–7^ These differences may arise from differing study populations, test kits, and procedures. We postulate that the settings in the rehabilitation center, which allowed for patients with mild symptoms to dwell in close quarters with other confirmed patients, may have brought about daily re-exposure and prolonged viral positivity durations.

Our study adds to the current knowledge concerning the COVID-19 outbreak and sheds light on the duration of RT-PCR positivity in young patients with mild symptoms. Seeing as young and healthy adults comprise much of the population workforce, the results of this study may assist in determining the isolation period for symptomatic adults and confirmed COVID-19 patients with mild symptoms.

Furthermore, we believe that achieving serial RT-PCR testing for all confirmed patients or outpatients with mild symptoms would require vast resource consumption. Therefore, achieving a further understanding of viral dynamics could help determine the correct temporality of testing for confirmed patients experiencing recovery.

It should be noted that negative RT-PCR testing does not necessarily dictate full clearance as both false-negative results^2,4,8^ and persistence of radiological abnormalities^9^ have been described. Additionally, the detection of SARS-CoV-2 in bodily fluids does not necessarily render an individual contagious.

Our study is limited by its small sample size and needs to be replicated on a mass scale to shed implications on the general population. It should also be noted that all patients in our cohort were symptomatic, and population screening efforts are required to determine the rate of disease and spread by asymptomatic individuals.

In conclusion, our study demonstrates that the median time for SARS-CoV-2 clearance is around three weeks in young adults with mild symptoms. These findings may provide evidence for instituting quarantine and return to work policies in settings primarily composed of young and healthy adults such as higher education institutions, high-schools and the military.

## Data Availability

Full patient data cannot be disclosed due to data security policies in the IDF. Deidentified data including patient clinical data is available per request.

## Acknowledgments

We would like to thank Eva Avramovich, MD, and Karina Castillo, BSN, of the Israel Defense Forces Medical Corps Public Health Branch for their assistance in data collection and approval of this manuscript. We would also like to thank Rayd Kayouf, RN, for his continued support in reviewing the manuscript and seeking approval for the collection and publication of the data involved.

